# Utility of human judgment ensembles during times of pandemic uncertainty: A case study during the COVID-19 Omicron BA.1 wave in the USA

**DOI:** 10.1101/2022.10.12.22280997

**Authors:** Srinivasan Venkatramanan, Juan Cambeiro, Tom Liptay, Bryan Lewis, Mark Orr, Gaia Dempsey, Alex Telionis, Justin Crow, Chris Barrett, Madhav Marathe

**Affiliations:** Biocomplexity Institute & Initiative, University of Virginia; Metaculus Inc.; Virginia Department of Health; Department of Computer Science, University of Virginia

## Abstract

Responding to a rapidly evolving pandemic like COVID-19 is challenging, and involves anticipating novel variants, vaccine uptake, and behavioral adaptations. Human judgment systems can complement computational models by providing valuable real-time forecasts. We report findings from a study conducted on Metaculus, a community forecasting platform, in partnership with the Virginia Department of Health, involving six rounds of forecasting during the Omicron BA.1 wave in the United States from November 2021 to March 2022. We received 8355 probabilistic predictions from 129 unique users across 60 questions pertaining to cases, hospitalizations, vaccine uptake, and peak/trough activity. We observed that the case forecasts performed on par with national multi-model ensembles and the vaccine uptake forecasts were more robust and accurate compared to baseline models. We also identified qualitative shifts in Omicron BA.1 wave prognosis during the surge phase, demonstrating rapid adaptation of such systems. Finally, we found that community estimates of variant characteristics such as growth rate and timing of dominance were in line with the scientific consensus. The observed accuracy, timeliness, and scope of such systems demonstrates the value of incorporating them into pandemic policymaking workflows.

---

The COVID-19 pandemic represents one of the most devastating pandemics in over a century, leading to more than 6 million deaths globally in the past two years. In addition to its huge toll on human life, it has led to an enormous economic and societal burden that will continue to impact the world long after the epidemiological effects subside. As of Spring 2022, there have been six prominent waves of infection at the national level in the United States, driven by various factors ranging from behavioral and policy changes, seasonality, and emergence of novel variants. The first Omicron wave, from November 2021 to March 2022, was caused primarily by the BA.1 sublineage of the Omicron variant (B.1.1.529). As seen in [1], this variant was the fastest until then to go from emergence to dominance (50% prevalence) to 100% prevalence, all within 5 weeks. Recent seroprevalence surveys [2] suggest that the BA.1 wave caused a significant number of infections, with nearly 25% of the population additionally infected (seroprevalence increase from 33.5% to 57.7% from December 2021 to February 2022), and up to a third of infected children aged 0-11 becoming newly seropositive in this period. This wave also led to sizable increases in at-home testing [3], event cancellations [4], and a renewal of widespread mask usage, in addition to other behavioral adaptations. While the Omicron BA.1 wave led to fewer deaths and hospitalizations per infection compared to the previous Delta wave, recent studies show that the variant could have been as deadly as prior waves upon adjusting for vaccinations, demographics, and comorbidities [5]. Furthermore, long-lasting physiological changes with poorly understood effects may remain even for those who have had mild cases.

Policy making during a pandemic is fraught with uncertainty in situational awareness (i.e., fog-of-war). Although epidemiological surveillance systems improved significantly during this pandemic, they still lag behind where they could be, given accessible, proven, and cost-effective capabilities that exist today. Further, recent advances in data science and availability of diverse streams of information (e.g., wastewater, genomics, case surveillance, search trends) allow us to assimilate them into a coherent picture and provide a reasonable prognosis [6]. Since early 2020, computational models have been influential in shaping a shared understanding of both facts on the ground and likely future scenarios within the scientific and policy communities, providing timely short-term forecasts and medium-term scenario-based projections for guiding policymakers and the general public. They have ranged from statistical and machine learning approaches that aim to exploit the patterns in the available data to mechanistic approaches that integrate available datasets with domain knowledge about their interactions and evolution. Multi-model ensembles have become the norm at the federal level [7, 8] for providing robust forecasts and projections. While the number of models range from 6-10 (for scenario-based projections) to more than 40 (for short-term forecasts), a rapidly spreading variant like Omicron requires quickly integrating emerging insights from preprints and adapting these models on-the-fly to reporting artifacts in the data, which is a significant undertaking.

In recent years, a complementary approach to model-driven epidemiological forecasting has emerged in the form of human judgment. Building upon the success of ‘wisdom-of-crowds’ approaches in other contexts [9, 10, 11], crowdsourced efforts for forecasting infectious disease outbreaks have been developed and have found reasonable success and adoption. Examples of early work in this area include [12, 13, 14]. [12] built an interactive platform for a small collection of disease modeling experts to ‘draw’ future trajectories of an ongoing influenza season, which are combined to provide short-term forecasts 1 to 4 weeks into the future. While this method performed among the best even amid computational models, the authors subsequently noted that this was difficult to scale and operationalize with the right incentives. [13, 14] have aimed to incorporate model-based forecasts and have allowed users to choose, rank, or combine them to create their forecast. A more detailed review of techniques prior to COVID-19 can be found in [15]. During the COVID-19 pandemic, this crowd-sourced approach has been used to evaluate vaccine policies [16] and combined with computational models to produce hybrid/ensemble forecasts [17, 18]. Recently, for the emerging monkeypox outbreak, human judgment forecasts were used to estimate cases, deaths, and impact across Europe, US and Canada [19].

In order to evaluate and effectively integrate such human judgment into real-time policy making, we conducted a Real-time Pandemic Decision Making (RPDM) tournament (https://www.metaculus.com/tournament/realtimepandemic/) in partnership with the Virginia Department of Health, on the community forecasting platform Metaculus. While other similar tournaments (https://www.metaculus.com/tournament/vdh/) focused on social, economic, and allied indicators over a longer horizon, the RPDM tournament focused on regularly renewed forecast questions pertaining to epidemic time series of reported cases, hospitalizations, and vaccine uptakes. Further, during the Omicron wave, a set of independent questions related to the pathogen’s characteristics and potential policy decisions were posted on the same platform (https://www.metaculus.com/questions/8759/forecasting-coronavirus-variant-omicron/).

In addition to individual forecasts, the platform generated a weighted ensemble (hereon referred to as the Metaculus Prediction). More details on the tournament structure, the questions, scoring rules, and ensemble generation are described in the Methods Section. In this paper, we primarily focus on the performance of the Metaculus prediction across various questions during the Omicron wave and summarize insights from a few key questions as part of the independent Omicron set. It is to be noted that the tournament structure and platform allowed for real-time updates to forecasts, as well as reformulation of the question set at different key points during the BA.1 wave, thus providing a unique lens into adaptation of human judgment systems during a rapidly evolving phase in the COVID-19 pandemic characterized by high uncertainty.

## Results

### Quantitative evaluation of forecasts

The forecasts were gathered through 6 tournament rounds spanning 18 weeks during November 2021 through March 2022. These rounds can be roughly grouped into two phases, of approximately 2 months each, namely: the surge phase (Rounds R1-R3 spanning Nov 12^th^, 2021 to Jan 14^th^, 2022), and the decline phase (Rounds R4-R6 spanning Jan 14^th^, 2022 to March 18^th^, 2022) of the Omicron wave. These two phases are divided by a central week-long peak period in cases and hospitalizations: the seven-day average of reported cases in Virginia peaked on January 13^th^, 2022, while hospitalizations peaked roughly a week later on January 21^st^, 2022. Some forecasting questions were modified between the phases to target the variables of interest better (see Table S1). Also, multiple forecast horizons were elicited for each variable to compare forecast uncertainty over time (see Table S2). Additionally, for variant characteristics, we use forecasts from a set of questions launched separately on the platform (see Data section in Methods).

Figures 1 and 2 show the performance metrics of these forecasts across targets and forecast horizons. Further, these metrics are captured at the beginning as well as the end of the forecast period (see Methods). As can be seen in case forecasts, they were mostly underestimated during the surge phase, whereas the decline phase was characterized by overestimates. This was especially true of longer forecast horizons (4wk ahead and 8wk ahead). While the performance generally improved over the forecast period, there were instances where the final estimates were less accurate than the early estimates (especially for 8wk ahead). At the time of closing, most vaccination forecasts were within a percentage point (pp.) of the ground truth, even in cases where they originally were off by up to 10 pp. For long-term case trends, the peak magnitude was mostly underestimated (during the surge phase), while the trough magnitude was mostly overestimated (during the decline phase). Further, as seen for R6 peak timing, the Metaculus prediction did not anticipate the surge due to subsequent Omicron variants (BA.2, BA.2.12.1), thus leading to the maximum error of 84 days (12 weeks).

**Figure 1:**
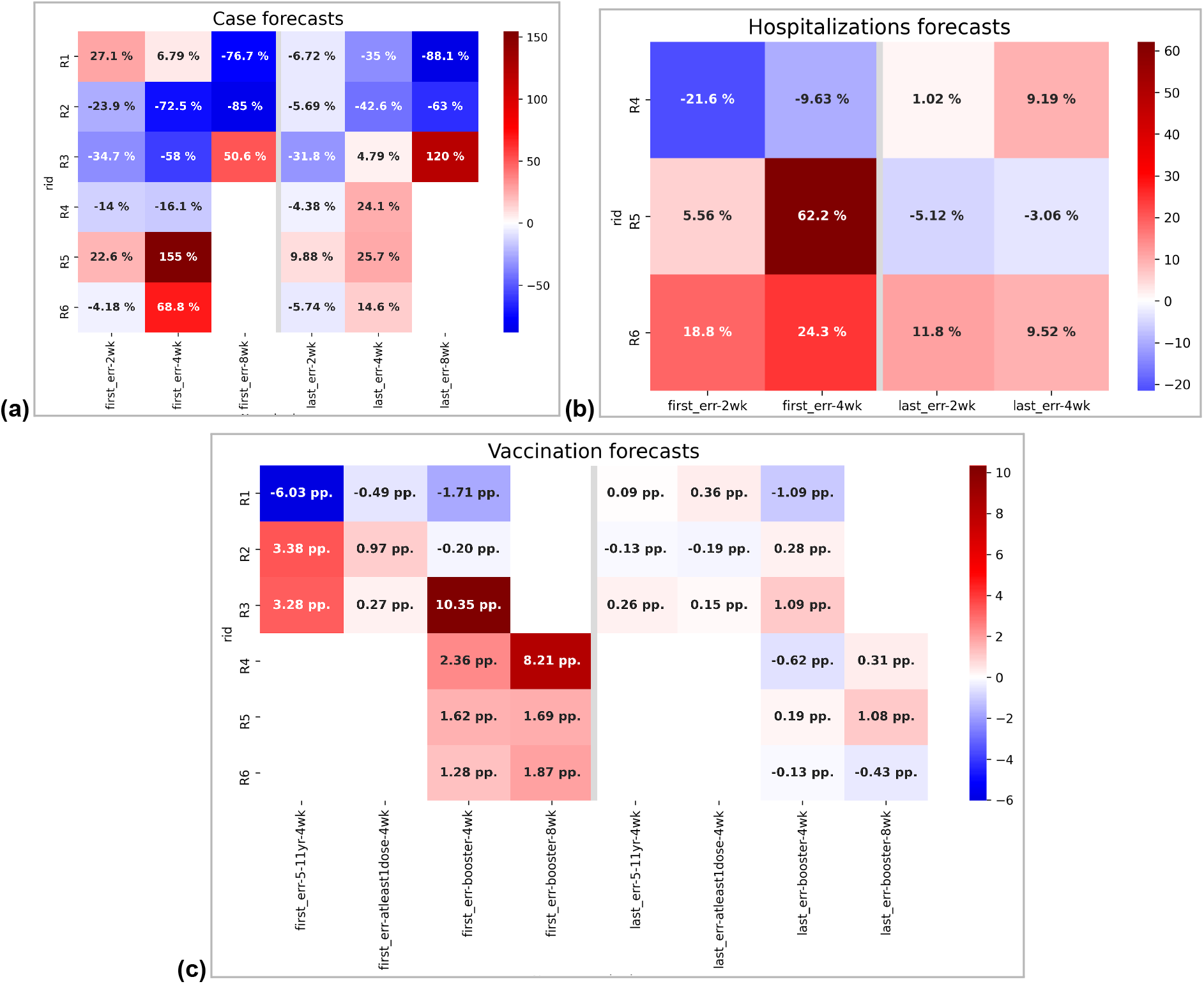
Performance metrics for case (%), hospitalization (%), vaccine forecasts (pp.) Underpredictions are shown in shades of blue, while overpredictions are shown in shades of red. The errors shown correspond to the first (day 1) and last (day 7 or 21 depending on the question) estimates during the open period. While case and hospitalization forecasts are summarized through percentage errors of the median, vaccination forecasts are summarized in percentage point difference. **(a)** Early rounds (R1-3) are characterized by case underpredictions, especially for the period around Omicron peak, whereas later rounds (R4-6) have overpredictions for both cases and hospitalizations **(b). (c)** Vaccine uptake, especially for boosters, is overestimated in the beginning but converges near the ground truth by the end of the forecast period. Comparison of case forecasts to those from multi-model ensemble is shown in Figure 3(a) & 3(b), while vaccine forecasts are compared to baseline models in Figure 4(c). Qualitative trajectories of the case and vaccination forecasts during R1-3 are available in Figures 5 & 6.

**Figure 2:**
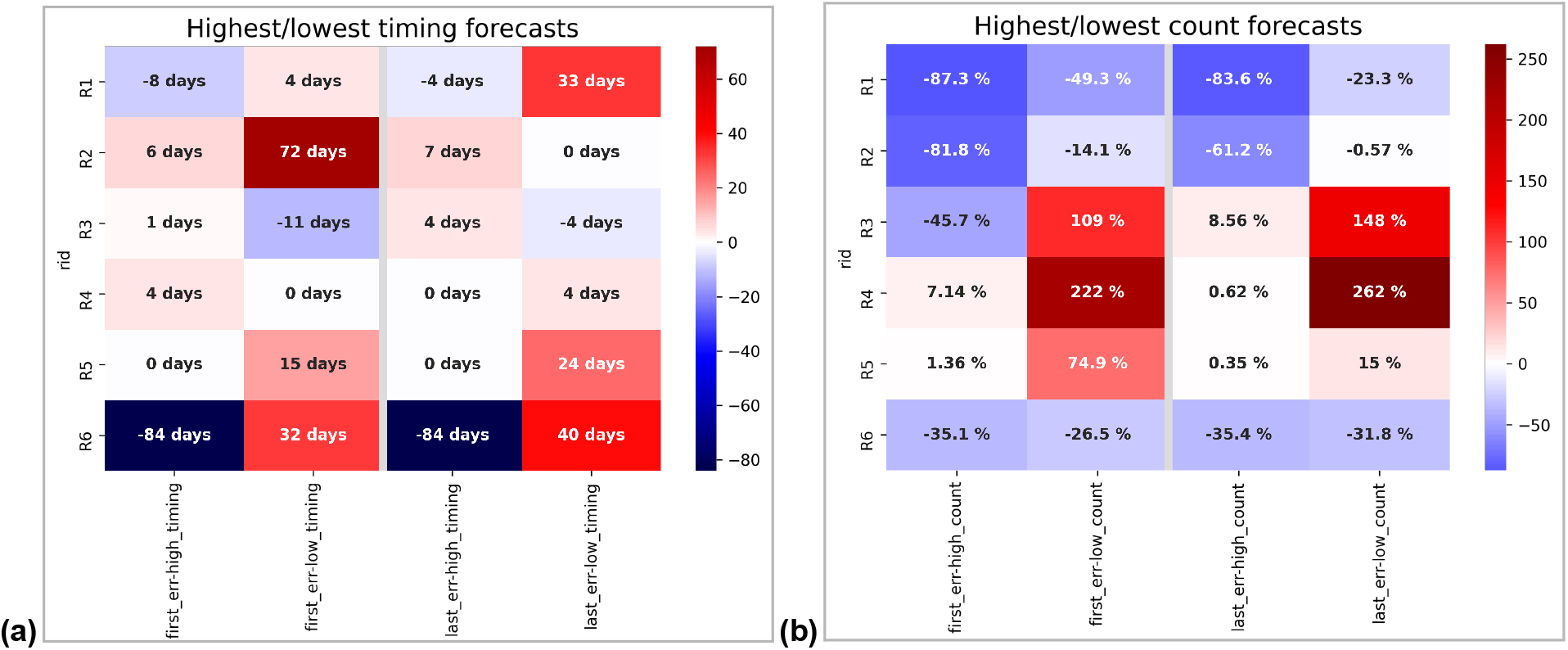
Performance of peak and trough forecasts (days, %) Underpredictions are shown in shades of blue, while overpredictions are shown in shades of red. The errors shown correspond to the first (day 1) and last (21) estimates during the open period. Note that peak and trough forecasts are sought for the trajectory over the next 12 weeks, hence the maximum error can be +/− 84 days. **(a)** Peak timing forecasts during the early rounds, especially for the BA.1 surge, were within approximately a week compared to ground truth. **(b)** Similarly, the trough timing forecasts during R2-4 were within a week. Peak magnitude forecasts during R1-3 are characterized by significant underestimation, while the trough magnitudes are overestimated during R3-5. Qualitative evolution of the peak and trough forecasts through rounds R1-6 are shown in Figure 7.

### Comparing case forecasts to multi-model ensemble

In order to further quantify the evolution of forecast performance over the forecast horizon, we utilize two metrics, namely *medMAPE* (*medMAE* for vaccine uptake) and *iqrCOV* (see Methods for details). As seen in Figure 3, these can be computed for each question across rounds and forecast horizons. The performance of the Metaculus prediction is then summarized by combining across forecasts at a given horizon, and compared with a similar metric for all forecasts from the unweighted ensemble of the CDC ForecastHub spanning the same period as the six rounds. While the Metaculus prediction is sampled at a daily resolution, the ForecastHub ensemble is only sampled at a weekly resolution thus we obtain performance metrics for forecast horizons corresponding to 7, 14, 21, 28 days.

**Figure 3:**
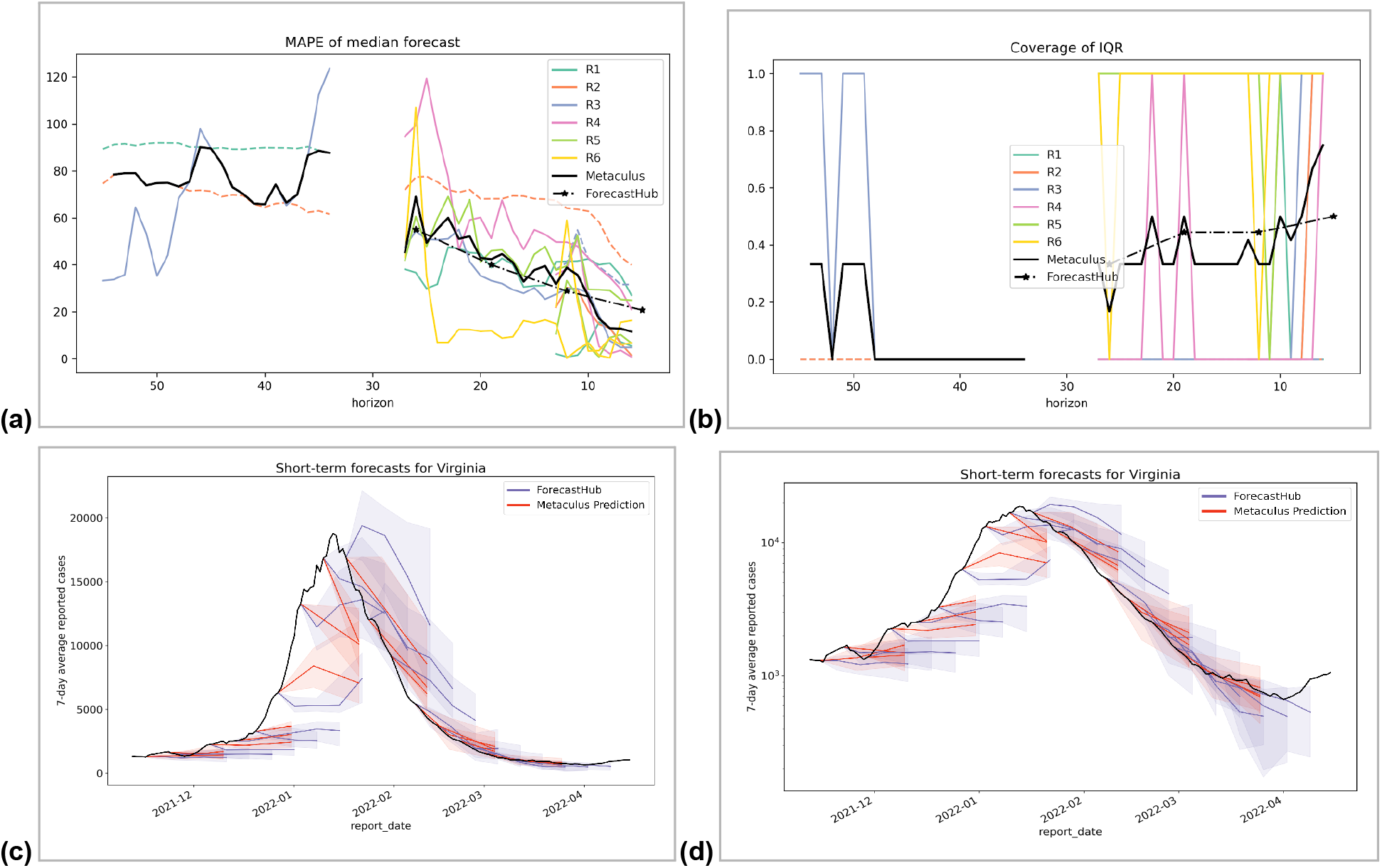
ForecastHub vs. Case forecasts Round 1-6. **(a)** ForecastHub forecasts have comparable MAPE to the median Metaculus prediction for the same forecast horizon across Rounds 1-6, except around 1-week ahead when Metaculus prediction is marginally better. **(b)** Similarly, the average coverage of IQR is generally comparable for the Metaculus prediction to the ForecastHub ensemble, with higher coverage around 1-week ahead. The forecasts are shown in **(c)** linear and **(d)** log scales to highlight the differences at peak and trough.

We note that the Metaculus prediction has comparable *medMAPE* to the ForecastHub ensemble through the entire window (from the forecast horizon of 28 days ahead). Further, the *iqrCOV* of the Metaculus prediction is comparable to that of ForecastHub during the same window, with marginally better performance for a forecast horizon less than 2 weeks. As seen in the visual comparison (also presented in log scale), while both approaches underestimated the Omicron surge, the Metaculus prediction has tighter uncertainty bounds post-peak.

### Comparing vaccine forecasts to linear baseline

As for vaccine uptake forecasts (Figure 4), we use medMAE to quantify the error in percentage points for each of the variables, and note that with an exception of 5-11 year-old uptake in R1, they are within 1% point of the ground truth with at least 3 weeks lead time. They also demonstrated higher IQR coverage (nearly 70%) through most of the forecast horizon. Further, to evaluate the utility of these forecasts, we compare them with linear baselines using different past regression windows (W = 3,5,7 days). Each of these models (labeled as ‘lincastW’ in Figure 4 where W is the regression window) perform linear regression on the recent W days of data and project forward with the same slope. We note that, in most instances, the Metaculus median prediction is more accurate at longer horizons than the linear estimate. Although there are instances where the linear model has a lower error, there is a good lagged correlation between the baseline and Metaculus prediction in trends.

**Figure 4:**
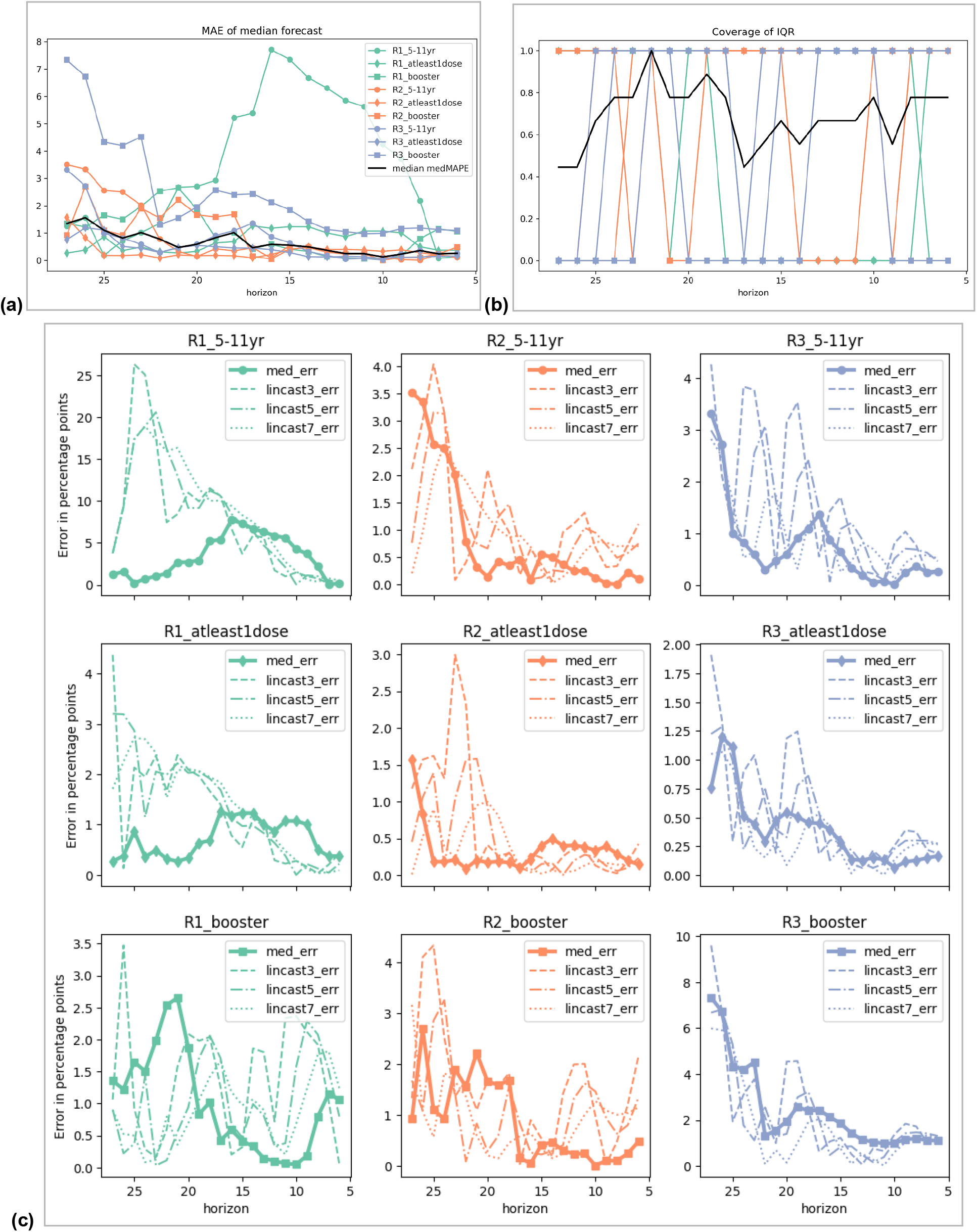
Vax forecast metrics: Round 1-3. **(a)** Except for 5-11 year-old vaccine uptake in the early phase during Round 1, others have consistent error performance, achieving within 1 percentage point of the target at least 3 weeks out. **(b)** The uncertainty intervals also seem to cover the ground truth on an average 70% of the time over the 4-week forecast horizon. **(c)** Compared to linear model baselines with different regression windows (3,5,7 days), the Metaculus median prediction is more robust and accurate for longer horizons.

### Qualitative patterns in case and vaccine forecasts

Even within the surge phase, forecasts during the three rounds were qualitatively different. R1 was characterized by limited knowledge of Omicron (except possibly some indicators outside the US). By R2, we saw a clear increase in infections as Omicron rose to dominance. This was further accelerated through R3, culminating in a peak of reported cases. Due to these differences in the information landscape over time, and heightened uncertainty during the surge, it makes sense to study these three rounds qualitatively in isolation.

From Figure 5, it is interesting to note that in R1, the short-term forecasts are well aligned with the latest ground truth, indicating limited trend signals being identified by forecasters. It is least influenced by Omicron, and the Metaculus prediction does not indicate seasonal effects would result in a new wave either (as seen in the 8wk ahead forecast). During R2, rapid changes in forecasts for 4wk and 8wk ahead can be seen, although it still matches the most recent ground truth (instead of projecting the exponential growth forward). It is worth noting that the users are limited by (and potentially influenced by) the range set as part of the question (indicated by the dashed blue line in Figure 1), and thus the upper bound limits some of these forecasts (see Table S1 and Table S3). In R3, even as the ground truth exceeded the upper bound, the Metaculus prediction was noticeably below it until the final few days. Finally, the 8wk ahead forecast in R3 can be seen as an early estimate of how long the Omicron wave may last, although predicting a slower decline than what eventually played out.

**Figure 5:**
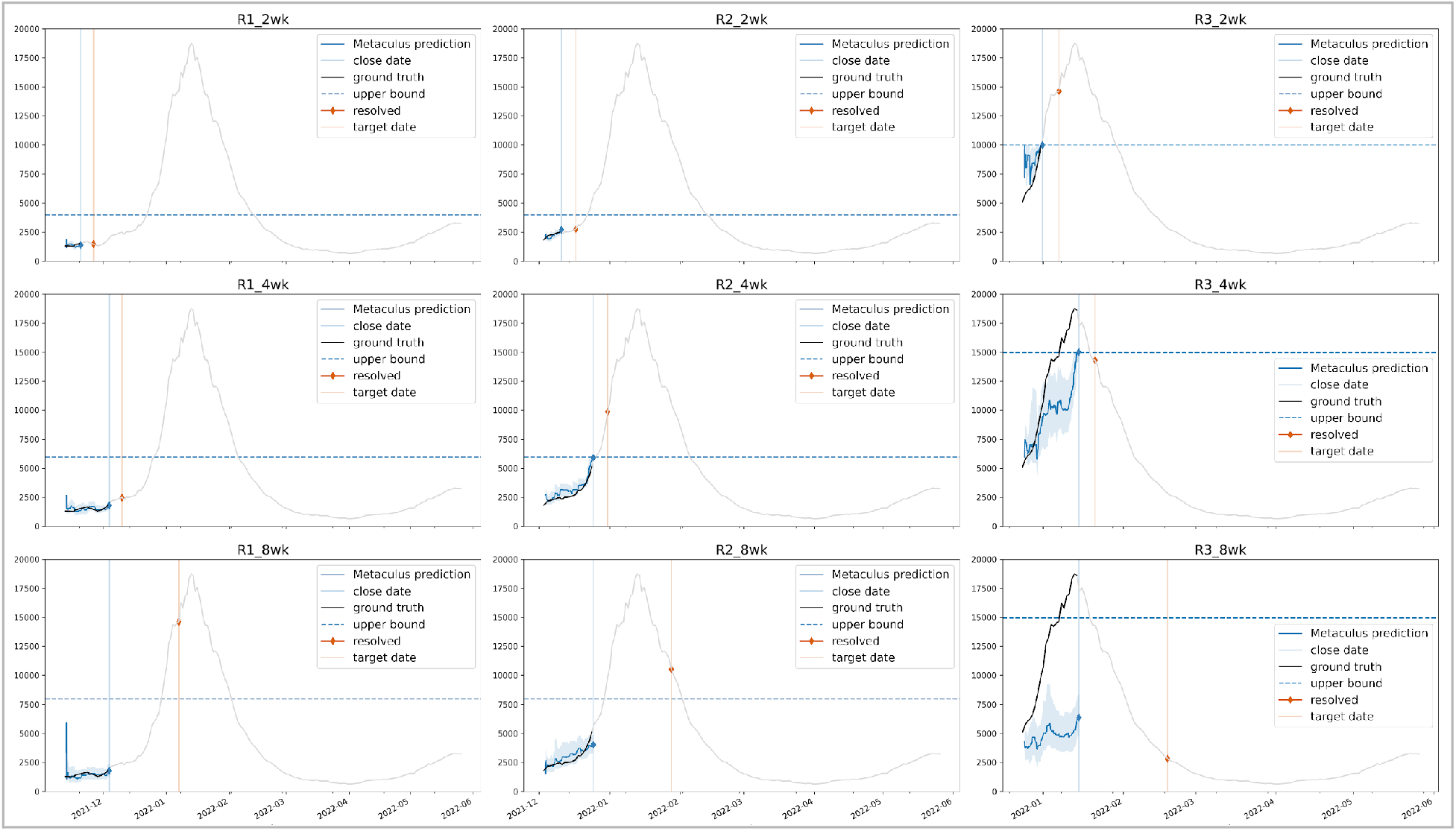
Case forecasts during the surge phase across different forecast horizons: Each column corresponds to a round (R1, R2, or R3), and each row corresponds to a forecast target horizon (2wk, 4wk, 8wk). Each panel shows the ground truth during the open period (black) and further extended beyond forecast horizon (gray). Metaculus predictions (blue) are shown with median and interquartile range during the open period for each question. The forecasts correspond to the time point denoted by the orange line. Final Metaculus prediction and the resolutions are shown as blue and orange diamonds. Dashed blue lines show the upper bound of the forecast range. While R1 (pre-Omicron) is characterized by forecasts tracking the ground truth, subsequent rounds show rapid adaptations as the surge becomes evident. Some of these forecasts (e.g., R3_2wk, R3_4wk) are concentrated around the upper bound. See Figure 1 for quantitative evaluation at the beginning and end of forecast period, and Figure 3 for comparison to the multi-model ensemble.

As for vaccine uptake forecasts (Figure 6), the surge phase covered various facets, ranging from rollout among 5-11 year olds, booster coverage, and overall full vaccination coverage. Since it corresponded to the early days in the rollout of vaccines for children under 12, R1 forecasts for uptake among 5-11 year-olds are more unstable. Across all these forecasts, the early estimates are optimistic, and a downward trend is observed as forecasts are continuously updated, and we approach the closing time for forecaster input. Some of these are also possibly influenced by the hidden period – the first 3/7^th^ of the forecast horizon of each question, during which the community median is not publicly displayed – and post-holiday effects, as seen in R3 booster coverage forecasts.

**Figure 6:**
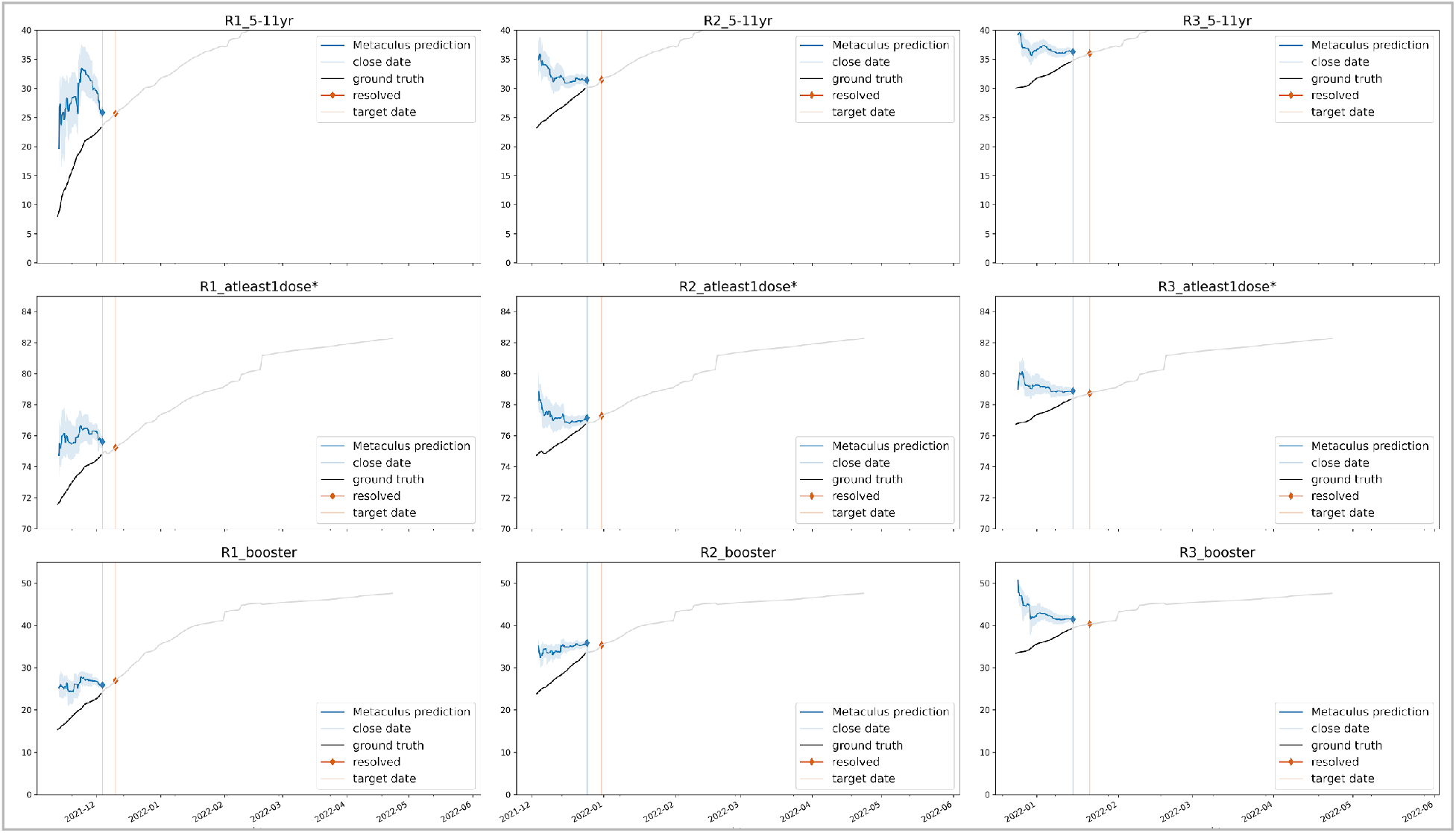
Vaccine forecasts during the surge phase across different forecast horizons: Each column corresponds to a round (R1, R2, or R3), and each row corresponds to a forecast target (5-11 year olds, at least 1 dose, boosters) at the one-month horizon. Each panel shows the ground truth during the open period (black) and further extended beyond forecast horizon (gray). Metaculus predictions (blue) are shown with median and interquartile range during the open period for each question. The forecasts correspond to the time point denoted by the orange line. Final Metaculus prediction and the resolutions are shown as blue and orange diamonds. Dashed blue lines show the upper bound of the forecast range. Most vaccine uptake forecasts begin as overestimates and subsequently are adjusted downward. This is especially evident for booster uptake forecasts during R3, with a nearly 10 percentage point drop over 3 weeks. Higher variability is seen in R1 5-11 year due to limited historical data. See Figure 1 for quantitative evaluation at the beginning and end of forecast period, and Figure 4 for comparison to baseline models.

### Qualitative trends in peak and trough forecasts

Long-term forecasts are inherently challenging to make due to changing behavioral variables (e.g., mask usage, social isolation, work-from-home, restricted travel) and policy changes. This is further accentuated during the emergence of novel variants since the exact magnitude and duration of their impact are unclear. We notice (Figure 7) that through the surge phase, this is reflected in the questions pertaining to peaks and troughs in the next 12 weeks for each round. While the peak timing forecast is set around mid-January from Round 1, this is more indicative of a historical estimate (from Winter 2020-21), hence the higher uncertainty. As Omicron emerges and surges, this estimate becomes tighter but does not change in median value. However, the effect of Omicron is observable through the increasing values for the peak magnitude forecast through Rounds 1-3. Likewise, while there is significant uncertainty regarding the ‘trough’ point in R1, by R2, the impact of the Omicron surge is evident, with most forecasters estimating the wave to last at least 12 weeks (until Feb 25^th^, 2022) from the start of R2 (Dec 3^rd^, 2021). It is also interesting that by R3, this estimate ‘switches’ to the right extreme of the forecast range, indicating that forecasters estimated the wave to be below the starting levels (on Dec 24^th^, 2021) in 12 weeks (by Mar 18^th^, 2022). With the benefit of hindsight, we now observe that both these forecasts were quite reasonable. During the decline phase, the peak forecasts were less informative, since they defaulted to the start of the forecast range (implying a continued decline from the Omicron wave). While this turned out to be true for rounds 4 and 5, for Round 6, due to the BA.2 wave, the true maximum was on May 20^th^, 2022, thus leading to an error of 84 days. One can also observe that the trough timings were overestimated for R5-6 (hinting at continued decline), while the magnitude forecasts were roughly aligned with the ground truth.

**Figure 7:**
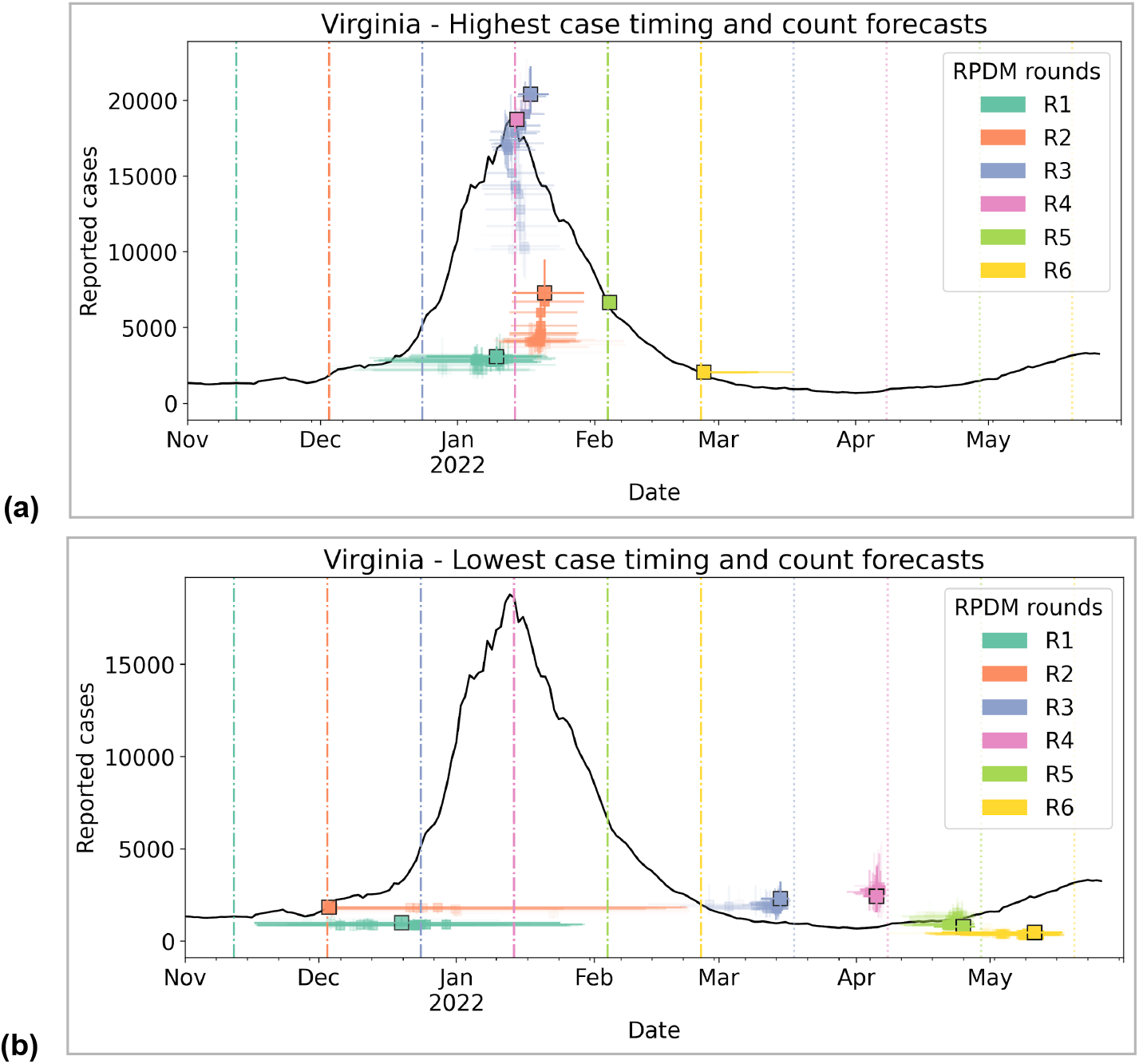
Comparison of peak/trough forecasts over R1-6. The median and interquartile ranges for **(a)** peak and trough timing and count forecasts are combined and tracked over the 21-day forecast period. The final forecasts from each round are shown with a black outline on the marker. Both peak and trough forecasts during the Omicron BA.1 surge phase are characterized by significant updates. During the decline phase, while peak forecasts default to the left end of the forecast range, the trough forecasts are reasonably consistent.

### Crowdsourced estimates for variant characteristics

Finally, Figure 8 shows that the value of such human judgment platforms go beyond just time-series forecasting. In both these cases, they are used to obtain timely best-guess estimates of variant characteristics and act as a crowdsourced curation of emerging literature. Figure 8(a) and 8(b) shows sharp adjustments in Omicron dominance time during December along with the underlying growth rate advantage relative to Delta. While initially forecast to be dominant only by mid-March 2022, the ensemble quickly converged in the latter half of December 2021, which matches now-known CDC estimates available online (https://covid.cdc.gov/covid-data-tracker/#variant-proportions) [Lambrou2022]. This is especially notable given that the CDC Nowcast estimates at the time were noisy —the estimate made on December 20 for the week of December 12-18 was 73.2% (95% CI: 34.0, 94.9) for nationwide Omicron prevalence. However, many forecasters on the platform quickly took issue with this and said that this estimate was implausible given the estimated growth rate of Omicron. Revised estimates for the week of December 12-18 later showed that Omicron prevalence for that week was much lower — with a point estimate of 22.5%, outside of the confidence interval of the previous estimate. Omicron indeed would not become the dominant variant until later in December. In this way, beyond just forecasting when Omicron would become dominant, the forecasters also served as a helpful check on the plausibility of modeled real-time estimates.

**Figure 8:**
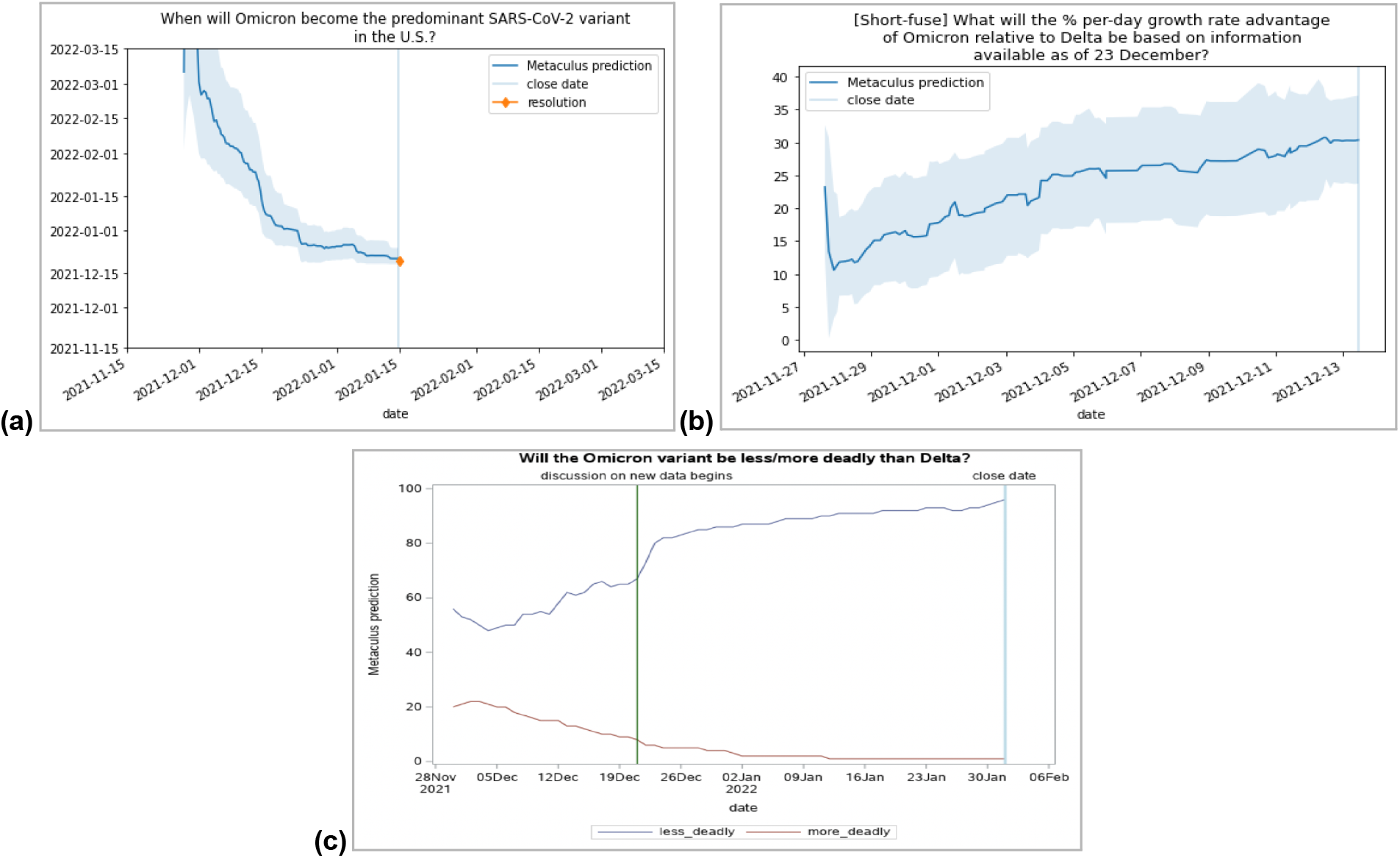
(a) Omicron prevalence forecasts; (b) Omicron growth rate forecasts; (c) Likelihood of Omicron severity. Early convergence on severity. Compare with literature. **(a)** After initial noisy estimates, the forecast for Omicron dominance timing steadily declines from mid-March to mid-December through the forecasting period. **(b)** This is also reflected in the estimated growth rate advantage, which steadily climbs from 10% to 30% during the forecast period. **(c)** Finally, the likelihood of relative severity shows significant updates around December 23^rd^ corresponding to new evidence becoming available through preprints.

Likewise, growth rate forecasts increased from almost 20% (nearly twice as fast as Delta over Alpha) to 30% (unprecedented in prior variants but observed in other countries like South Africa, Denmark, and Belgium) over the forecasting period from late November to mid-December. This estimate of the growth rate advantage of Omicron over Delta was a real-time attempt to estimate this key parameter to inform the answers to other questions (e.g., when Omicron would overtake Delta as the dominant variant). This question was purposefully set up to resolve soon after the forecasting period and preclude the possibility of other sub-variants of Omicron influencing the growth rate estimate. Notably, we experimented with lax resolution criteria in which pre-prints and other non-peer-reviewed analyses would count toward a resolution so long as the authors were known to be reliable, which indicates substantial flexibility in approaches to human judgmental forecasting. The final resolution value of 37% was higher than the final Metaculus prediction of 30% but still within the IQR.

Finally, we also note the quick emergence of consensus concerning Omicron’s relative severity over Delta (Figure 8c). At the time Omicron emerged, reports from South Africa indicated that on a per-case basis, the severity of Omicron was lower than that of Delta. However, it was difficult for researchers and public health officials to quantify the likelihood of this being true. By December 6, Metaculus forecasters consistently assigned a greater than 50% chance of Omicron being less lethal than Delta and a 20% or lower chance of it being more lethal than Delta. Forecasters gradually updated until they sharply adjusted their forecasts for Omicron being less lethal from 67% on December 21 to 80% on December 23. This is likely attributable to two pre-prints being released on December 22, which both found Omicron is associated with a reduction in the risk of COVID-19 hospitalization when compared to Delta [20, 21], which prompted discussion on the relevant Metaculus question (https://www.metaculus.com/questions/8766/omicron-variant-less-deadly-than-delta/#comment-76033). By December 24, forecasters were consistently assigning a greater than 80% chance to this being the case. The rapid emergence of this consensus on the relative severity of the disease due to the Omicron variant could have played a pivotal role in public health decision-making.

## Discussion

The Omicron BA.1 wave highlighted the rapidity with which novel variants or emerging pathogens can sweep through a population even with sufficient prior immunity (in this case through previous waves of infection and vaccinations). Decision-making under such settings is time-sensitive, but there is scant evidence for informing such decisions. While computational models can be repurposed to accommodate available data, robust parameter estimates can only be obtained through careful coordination and deliberation across multiple groups of scientists. This pilot study demonstrates that human judgment ensembles provide valuable signals for real-time pandemic decision-making during such high uncertainty. In addition to forecasts of comparable accuracy to that of computational models, such systems could provide timely updates to a diverse set of questions. Further, with the decision maker in the loop, such systems allowed for iterations on the question set, thus focusing on the appropriate variables of interest during different phases.

It has to be noted that such human judgment systems do not exist in a vacuum. They operate in an environment with widely available and well-maintained disease surveillance dashboards, publicly available model forecasts, active social and news media discussion, and rapid dissemination of scientific findings via preprints. The human participants then serve as effective information aggregators who can produce forecasts for variables of interest through mental models. Platforms such as Metaculus are essential for the effective collection, tracking, and ensembling of such forecasts to be useful for policymakers.

The pilot study is not without limitations. Experiment design is a time-consuming process that involves careful choice of questions, exact phrasing, forecast range, cadence, and resolution criteria. Further, there is limited understanding on how and when individual users update forecasts, and whether they ensure consistency across questions. Linking the user updates to changes in the information environment is crucial to filter out spurious changes, and interpret such ensembles. While comments and discussion can be solicited, a more formal mechanism for specifying their mental models and data inputs is needed, much like the metadata associated with computational models. Further, understanding the causal linkages between the variables of interest will allow better sampling of the information space, and enable soliciting the most relevant inputs at an appropriate frequency from the forecasters to minimize their cognitive load. Better ensembles can also be constructed by exploiting similarities in user forecasts and expertise (e.g., domain scientists, quantitative analysts, regular citizens) across questions.

We believe that the utility of such systems go beyond providing direct forecasts. One must consider combining them with computational models to make the most of such ensembles. Since they can aggregate information available via preprints on parameter estimates, they can subsequently be used in computational models to provide projections. Forecast information could also provide ‘public pulse’ estimates of behavioral aspects such as mask use and vaccine uptake, which can augment traditional survey-based and participatory surveillance mechanisms. Further, forecasts from such systems can be included in a statistical ensemble (much like the ForecastHub) along with other model-based forecasts to provide robust inputs for the policymaker. Human forecasters can be aided by interactive computational models and other analytical tools to help assimilate domain knowledge and minimize certain cognitive biases. Finally, experiment design can be significantly improved by focusing on data gaps in the surveillance or the computational models that can benefit from such a system. Exploring the role of policy decisions through conditional questions will allow for anticipating system dynamics and provide more valuable forecasts.

## Methods

### Data

Human judgment forecasts were compared against ground truth datasets compiled by the Virginia Department of Health (reported cases, vaccine uptake) and the Virginia Hospital & Healthcare Association (for current hospitalizations). These were obtained from the Virginia Open Data Portal (see Data Availability Statement). Fully vaccinated coverage is defined as the fraction of individuals with two doses of a two-dose vaccine or one dose of a single-dose vaccine, with respect to the overall population. A similar calculation was performed for booster coverage, although the federal doses were not included as part of the numerator (see Vaccine Summary Dashboard https://www.vdh.virginia.gov/coronavirus/see-the-numbers/covid-19-in-virginia/covid-19-vaccine-summary/ for details). Multi-model ensemble forecasts generated as part of the CDC ForecastHub effort were obtained from the Github repository (https://github.com/reichlab/covid19-forecast-hub) spanning the period corresponding to the six rounds. Ensemble forecasts from Metaculus were obtained through the public API for questions in the Real-Time Pandemic Decision Making tournament (https://www.metaculus.com/tournament/realtimepandemic/) and the set of independent questions pertaining to the Omicron variant (https://www.metaculus.com/questions/8759/forecasting-coronavirus-variant-omicron/).

### Tournament Structure and Interface

The forecasting exercise was conducted as part of the Real-Time Pandemic Decision Making tournament on the Metaculus platform. It was conducted over six rounds, starting November 12^th^, 2022, and concluding on March 18^th^, 2022. Each round had 10-12 questions and was open for 3 weeks (with the exception of questions with shorter forecast horizons). Rounds did not overlap, i.e., when questions for one round closed for forecasting, a new round of questions opened. More information is provided in the Supplementary Material. Details on the questions and forecast horizons are provided in Table S1. Start and close times for each round, along with exact dates for forecast horizons are provided in Table S2.

Each question is provided with relevant background context, and an unambiguous question with the target date and metric of interest. The resolution criteria are also clearly specified with the source (dashboards, in this case) and the date when the resolution will be made (allowing for data stability). Forecasters provide probabilistic forecasts by constructing a mixture of logistic distributions over the support set specified as part of the question. For binary forecasts, individuals report their forecasts as a probability. Forecasts can be updated at any point during the question’s open period. To encourage independent forecasts, forecasters could not see each others’ forecasts or the community forecast for the first 3/7th (~43%) of the open period of a question. For the remaining period, the community forecast (pointwise median among available forecasts with a weighting scheme prioritizing recent forecasts) is made visible. Finally, while making/updating the forecasts, in addition to the community ensemble, users are also shown a real-time estimate of current points corresponding to their forecast, depending on the community prediction and the ultimate result.

We received a total of 8355 probabilistic predictions from 129 unique users across all the 60 questions. On average, there were about 25-30 unique users per question and about 100-200 predictions per question. The number of users and predictions peaked around R3 (see Figure S1).

### Forecast generation

A forecaster on the Metaculus platform can submit a predictive density *f* as a convex combination of up to five logistic distributions

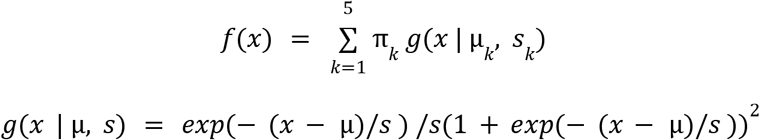

where *π*_*k*_ is a nonnegative weight associated with the kth logistic distribution *g*(*x*|*μ*_*k*_, *s*_*k*_) and all weights are constrained to sum to one. By default, a forecaster is presented with a single logistic density and a slider bar underneath this density that contains a square and two circles. A forecaster can change the value of *µ* by shifting the square to the left for lower values and to the right for higher values. The parameter *s* is adjusted by independently sliding the circles left and right, thus leading to a generalized asymmetric distribution. Users can add up to 4 additional components, along with sliders to control the weights for each component. A user’s predictive density is continually updated in the browser to facilitate building their forecast for submission. When the forecaster is satisfied with their prediction, they press “submit”. After the first submission, forecasters may revise their original prediction as often as they choose.

Forecasters construct a predictive density over a bounded interval that is presented on either a linear scale or a log scale (base 10). The choice of interval and linear vs. log scale is made by question developers at Metaculus. Once 10 predictions from 10 unique forecasters are submitted to the Metaculus platform, an ensemble of these predictions is revealed to the community after the hidden period. An ensemble predictive density *f* was created as a weighted combination of all *M* individual forecaster densities

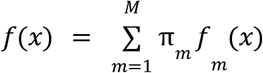

where the weight assigned to forecaster *m*, annotated as π_*m*_, is a function of the forecaster’s accuracy on past questions where they submitted a prediction with ground truth available and how recently they submitted a prediction. This ensemble strategy applies to questions that ask a forecaster to submit a predictive density over a closed interval.

### Scoring and Tournament Leaderboard

Metaculus scoring rules are built on the logarithmic score, a commonly used metric for probabilistic forecasts. We begin with the description for a binary question and show its generalization for continuous questions. For a single binary forecast of probability *p*, this corresponds to the natural logarithm of the probability assigned to the outcome, i.e., *S = ln*(*p*) if the event occurred, and *S = ln*(1 − *p*) if it did not. To measure the accuracy of a user’s forecast probability *p* relative to the community forecast probability *cp*, relative log score for a binary question is defined as *ln*(*p*/*cp*) if the event occurred and *ln*(1 − *p*/*cp*) if it did not occur. For continuous questions, *p* is the density of user forecast probability distribution at the resolution value and *cp* is the density of the community probability distribution at the resolution value.

Question score is a time average of a user’s relative log score over the duration of a question (also known as the open period). For periods when the user was not active (i.e., had not yet made a forecast), the relative log score is set to 0 (i.e., *p = cp*). Question coverage is the percentage of the relevant period for which a user has an active forecast. Since community predictions are hidden in the beginning (for 3/7th of the open period), forecasters must rely on their independent judgment. A forecaster’s question score is the time average of their relative log score over the duration of a question.. A user’s tournament score is then the total of their individual question scores, and the tournament coverage is calculated as the average of their question coverages. The tournament take, i.e., share of the ultimate prize pool, is defined as the tournament coverage times the exponential of their tournament score. A real-time leaderboard for all forecasters is made available through the tournament landing page.

### Metaculus Prediction

While the user’s tournament scores are obtained in relation to a community median ensemble, Metaculus also provides a trained ensemble which, in addition to weighting forecasts for their recency, also accounts for the individual forecaster’s track record across the entire platform.

### Forecast Evaluation metrics

We define two separate metrics to characterize the performance of the Metaculus prediction and compare them against the multi-model ForecastHub ensemble. *medMAPE* is the absolute percentage error between the median of the Metaculus (or ForecastHub) ensemble and the observed ground truth. *iqrCOV* is an indicator (0 or 1) of whether the actual ground truth falls within the interquartile (25th to 75th percentile) of the Metaculus (or ForecastHub) ensemble. Given the ground truth *y*_*t*_ for a target of interest, and *ŷ*_*d,p*_ the *p*^th^ percentile of the Metaculus (or ForecastHub) forecast *d* days ahead:

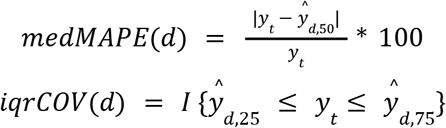

Since these are available across different timepoints, we group them by forecast horizon (the duration from forecast date to target date) and compute the median value for *medMAPE* and average for *iqrCOV*. Note that, ForecastHub forecasts are made on a weekly cadence for 1, 2, 3, 4 *epiweeks* (Sunday-Saturday) ahead and are submitted on the Monday of the first *epiweek* ahead, whereas Metaculus case forecasts are solicited for the 7-day moving average (right aligned) on a Friday. For equivalent comparison, ForecastHub forecasts are converted to 7-day moving average on Saturdays (right aligned) before evaluation. Since vaccine uptake forecasts are already in percentages, hence *medMAPE* is replaced by *medMAE* (mean absolute error of median of the Metaculus prediction) and reported in percentage points.

### Ethics Statement

No identifiable information was collected, and no additional recruiting was done for this study beyond the users who were already part of the Metaculus platform. Users were incentivized only through the overall prize pool, and were not individually contacted. The study was approved by The University of Virginia Institutional Review Board for Social & Behavioral Sciences.

### Data Availability Statement

Metaculus predictions for all the RPDM questions are available for public viewing on the platform (https://www.metaculus.com/tournament/realtimepandemic/). Further, the cleaned data used for analyses is available at (https://github.com/NSSAC/metaculus-rpdm-data). Cases, hospitalizations, and vaccination data were obtained from the public datasets maintained by the Virginia Department of Health (https://data.virginia.gov/Government/VDH-COVID-19-PublicUseDataset-Cases/bre9-aqqr, https://data.virginia.gov/Government/VDH-COVID-19-PublicUseDataset-KeyMeasures-Hospital/28wk-762y, https://data.virginia.gov/Government/VDH-COVID-19-PublicUseDataset-Vaccines-DosesAdmini/28k2-x2rj). Forecasts from COVID-19 Forecast Hub were obtained from their Github repository (https://github.com/reichlab/covid19-forecast-hub)

## Acknowledgments

The authors would like to thank the community forecasters for their participation and contributed predictions throughout the tournament, as well as the team at Metaculus for efficient and smooth operation of the tournament. The authors also acknowledge the scientific feedback from members of the Biocomplexity Institute COVID-19 Response Team at University of Virginia. This work is partially supported by the Virginia Department of Health (COVID-19 Modeling Program VDH-21-501-0135), National Science Foundation (Collaborative Research: Expeditions: Global Pervasive Computational Epidemiology CCF-1918656), and University of Virginia (Strategic Investment Fund SIF160).

