## Supplemental Information for "Utility of human judgment ensembles during times of pandemic uncertainty: A case study during the COVID-19 Omicron BA.1 wave in the USA"

[2]*Metaculus Inc.*

**Table S1: Questions in each tournament round**

| | Round - Forecast Horizon | | | | | |
|---|---|---|---|---|---|---|
| **Target** | **R1** | **R2** | **R3** | **R4** | **R5** | **R6** |
| 7-day avg. reported cases | 2,4,8* wk | 2,4*,8* wk | 2*,4,8 wk | 2,4 wk | 2,4 wk | 2,4 wk |
| Fully vaccinated coverage (%) | 4wk | 4wk | 4wk | X | X | X |
| 5-11yr old vaccine coverage (%) | 4wk | 4wk | 4wk | X | X | X |
| Booster coverage (%) | 4wk | 4wk | 4wk | 4,8 wk | 4,8 wk | 4,8 wk |
| 7-day avg. current hospitalizations | X | X | X | 2,4 wk | 2,4 wk | 2,4 wk |
| Highest 7-day avg cases - timing | 12wk | 12wk | 12wk | 12wk | 12wk | 12wk |
| Highest 7-day avg cases - count | 12wk* | 12wk* | 12wk | 12wk | 12wk | 12wk |
| Lowest 7-day avg cases - timing | 12wk | 12wk | 12wk | 12wk | 12wk | 12wk |
| Lowest 7-day avg cases - count | 12wk | 12wk | 12wk | 12wk | 12wk | 12wk |

*case targets for R1(8wk), R2(4wk, 8wk), R3(2wk), and peak targets for R1,R2 were out of range (see Table S3).

**Table S2: Time windows and horizons corresponding to each round**

| R# | Start time* | Close time† | 2 wk | 4wk | 8wk | 12wk |
|---|---|---|---|---|---|---|
| **1** | Nov 12th, 2021 | Dec 3rd, 2021 | Nov 26th, 2021 | Dec 10th, 2021 | Jan 7th, 2022 | Feb 4th, 2022 |
| **2** | Dec 3rd, 2021 | Dec 24th, 2021 | Dec 17th, 2021 | Dec 31st, 2021 | Jan 28th, 2022 | Feb 25th, 2022 |
| **3** | Dec 24th, 2021 | Jan 14th, 2022 | Jan 7th, 2022 | Jan 21st, 2022 | Feb 18th, 2022 | Mar 18th, 2022 |
| **4** | Jan 14th, 2022 | Feb 4th, 2022 | Jan 28th, 2022 | Feb 11th, 2022 | Mar 11th, 2022 | Apr 8th, 2022 |
| **5** | Feb 4th, 2022 | Feb 25th, 2022 | Feb 18th, 2022 | Mar 4th, 2022 | Apr 1st, 2022 | Apr 29th, 2022 |
| **6** | Feb 25th, 2022 | Mar 18th, 2022 | Mar 11th, 2022 | Mar 25th, 2022 | Apr 22nd, 2022 | May 20th, 2022 |

*Start and Close times are Friday 12PM EST. Each round lasted three weeks.
†2wk ahead questions are closed 1 week from start time

**Table S3: Case forecast questions with ground truth above upper bound.**

| short_code | Upper bound | Ground truth |
|---|---|---|
| **R1_8wk** | 8000 | 14645 |
| **R2_4wk** | 6000 | 9883 |
| **R2_8wk** | 8000 | 10556 |
| **R3_2wk** | 10000 | 14645 |
| **R1_peak_12wk** | 10000 | 18782 |
| **R2_peak_12wk** | 10000 | 18782 |

**Figure S1: Number of unique users and predictions across rounds**

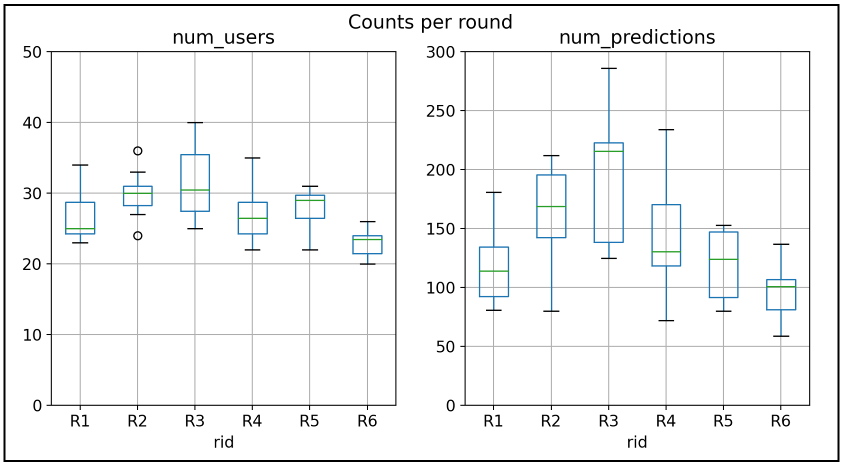
